# Smoking drives an epigenetic memory of aberrant hematopoiesis

**DOI:** 10.64898/2026.05.14.26353250

**Authors:** Charles E. Breeze, Gabriel A. Goodney, Hantao Wang, Aubrey K. Hubbard, Jungeun Lim, Mitchell J. Machiela, Thanh T. Hoang, Marie Richards-Barber, Christine Tran, Matthew Tolentino, Mark Hansen, Rishi Porecha, Nicole Renke, Wanding Zhou, Nora Franceschini, Sonja Berndt, Jonathan Hofmann, Mikyeong Lee, Stephanie J. London, Jason Y.Y. Wong

**Affiliations:** Occupational and Environmental Epidemiology Branch, Division of Cancer Epidemiology and Genetics, National Cancer Institute, National Institutes of Health, 9609 Medical Center Drive, Rockville, MD 20850, USA; Epidemiology and Community Health Branch, Division of Intramural Research, National Heart Lung and Blood Institute, National Institutes of Health, 10 Center Drive, Bethesda, MD 20892, USA; Epidemiology Branch, National Institute of Environmental Health Sciences, National Institutes of Health, Research Triangle Park, NC, USA; DLH LLC, Bethesda, MD, USA; Department of Pediatrics, Division of Hematology-Oncology, Baylor College of Medicine, Houston, Texas, USA; Cancer and Hematology Center, Texas Children’s Hospital, Houston, Texas 77030, USA; Westat, Inc., Durham, North Carolina, United States of America; Illumina, Inc., San Diego, CA 92122, USA; Center for Computational and Genomic Medicine, The Children’s Hospital of Philadelphia, Philadelphia, PA 19104, USA; Department of Pathology and Laboratory Medicine, University of Pennsylvania, Philadelphia, PA 19104, USA; Department of Epidemiology, University of North Carolina, Chapel Hill, NC, USA

## Abstract

Tobacco smoking induces DNA methylation (DNAm) changes in blood and other tissues, which may influence chronic health outcomes. However, the breadth of smoking-related DNAm changes remains unmapped, offering a space for employing novel technologies. To expand our understanding of smoking impacts on DNAm, we conducted an epigenome-wide association study (EWAS) comparing ever smokers to never smokers, using blood from a multiethnic U.S. study population (n=887). We employed the newly developed Illumina Methylation Screening Array (MSA) covering 269,094 unique sites, including 123,776 CpGs not assayed in previous EWAS. Trans-ethnic meta-analysis identified 152 differentially methylated positions (DMPs) associated with ever-smoking status (n=764); European-specific analysis yielded 129 DMPs (n=674), including 106 overlapping with trans-ethnic analysis. A separate, large-scale replication EWAS (n=2,190) confirmed 91 trans-ethnic and 77 European-specific DMPs. Among our findings, we identified 61 DMPs at CpGs novel to the MSA platform, including near both new and known smoking-associated genes. Most notably, we uncovered a dense cluster of 12 DMPs within a 1117 bp region of *ECEL1P1*, forming the most long-lasting, persistent smoking-associated DMR ever detected, even among former smokers who quit decades prior. We also detected new signals at *AHRR*, a well-known locus for smoking-related DNAm changes. eFORGE analysis revealed that detected smoking-associated DNAm changes are predominantly located in hematopoietic stem and progenitor cell (HSPC) DNase I hotspots, aligning with gene set enrichment analyses that highlighted pathways related to hematopoietic stem cell differentiation. Our findings suggest that HSPCs serve as a reservoir for an epigenetic memory of smoking. Additionally, we observed short-term cell-specific smoking-associated DNAm changes in myeloid cells. Our results demonstrate the utility of the MSA in expanding our knowledge of both transient and persistent environmental exposure-associated DNAm changes.

**Highlights:**

- Applied the state-of-the-art Methylation Screening Array (MSA), 269,094 unique sites including 123,776 not studied previously, which were selected for likely functional relevance.
- Identified 61 novel smoking-associated differentially methylated positions (DMPs), annotated to novel genes as well as genes previously associated with smoking-related DNA methylation.
- Smoking-associated DMPs were enriched in regulatory elements of hematopoietic stem and progenitor cells (HSPCs, via eFORGE analyses and GSEA) and HSPC regulatory genes (e.g. *RUNX1*), implicating HSPCs as reservoirs of long-term epigenetic memory.
- 12 DMPs collectively form the most long-lasting, persistent smoking-associated differentially methylated region (DMR) detected so far, spanning a 1117 bp region at *ECEL1P1*.
- Smoking drives two distinct classes of DNAm alterations: transient, myeloid-specific changes and persistent, cell-type-shared signatures originating in HSPCs, forming a dual-track model of smoking-induced epigenetic remodeling.

## Introduction

Tobacco smoking is one of the strongest factors driving alterations in DNA methylation (DNAm) in epigenome-wide association studies (EWAS)^1–4^. Multiple EWAS have identified a broad spectrum of DNAm changes in blood associated with smoking, with some of the most robust and reproducible signals observed at loci such as the aryl hydrocarbon receptor repressor (*AHRR*) locus. Remarkably, these smoking-induced epigenetic modifications persist for several years after smoking cessation and are detectable across a variety of tissues. The magnitude of these effects is detectable at the level of cell type–specific DNAm changes^5–7^, which have demonstrated associations with smoking in discrete cell populations, most notably in myeloid cells^7^. Independently, there are reports about shifts in the relative count of different cell populations during smoking. Several reports indicate that smokers display an increased proportion of monocytes in peripheral blood^8,9^. A better mechanistic understanding of smoking-associated DNAm changes is critical, as these patterns may mediate the chronic health effects of long-term tobacco use. Current characterization of smoking-related DNAm changes remains limited, and shows great potential for improvement with expanded coverage of the genomic CpGs assayed. The laboratory methods used to assay DNAm necessarily influence our understanding of smoking-related DNAm changes. Most smoking EWAS to date have used Illumina Infinium arrays, which balance cost, genomic coverage and DNAm measurement precision^10,11^. One of the caveats of previous Illumina EPIC and 450k array-based EWAS is their inclusion of many CpG sites that exhibit limited DNAm changes in blood, potentially overlooking more informative regions of interindividual variation^12^. Addressing this limitation, novel DNAm technologies such as the recently developed Illumina Methylation Screening Array (MSA) have emerged^13^. The MSA was developed as a more compact, scalable and focused array while retaining the measurement accuracy of existing platforms. Crucially, the MSA targets a wide range of novel CpG sites with increased variance, including novel regulatory elements and correlated regions of systemic interindividual variation (CoRSIVs)^12^, aiming to capture dynamic regions of the DNA methylome that were previously uncharacterized in order to broaden biological insight. In addition, this array also includes a large number of CpGs from previous arrays, focusing on sites with established associations across a range of exposures and traits—such as smoking, birth weight and aging—as well as other significant epigenetic markers^13^, allowing for a direct comparison of MSA with EPIC and 450K array measurements^13^.

In this study, our overarching aim was to discover novel smoking-related differentially methylated positions (DMPs) and genes in blood to expand our mechanistic understanding of the effects of current and past smoking, as well as validate and refine previously reported DNAm changes. Importantly, we aimed to integrate these findings with other genomic data to elucidate the underlying mechanisms that contribute to the long-term persistence of smoking-associated epigenetic alterations and their impact on cellular function. To accomplish these aims, we applied the recently developed Illumina MSA platform to blood samples collected from 887 participants from the Agricultural Lung Health Study (ALHS), a multiethnic case-control study of asthma nested within a large prospective cohort study in the United States. To replicate our results in current vs. never smokers, we used data from a published blood-based EPIC array EWAS meta-analysis across 5 cohorts including 2560 current smokers and 8521 never smokers^4^. To replicate our results in former vs. never smokers, we performed a similar analysis of 667 former smokers compared to 1523 never smokers using the EPIC array in a larger sample of the ALHS.

## Results

### Novel smoking-associated methylation genes and CpGs identified using the MSA

We performed an EWAS of smoking status using the Illumina Infinium MSA. Analyses contrasting current versus never smokers, and former smokers versus never smokers, revealed 61 new DMPs (Bonferroni-adjusted significance threshold of p≤2.17×10^−7^ corresponding to 0.05/230,325 MSA probes, **Figure 1b**, **Tables S1-S8**). Importantly, the broadest regional variation detected with the MSA was a cluster of 12 correlated CpGs within the *ECEL1P1* gene, with cg04284215 and cg04284216 among the strongest signals. We found additional novel loci at *CNTNAP2* (cg11855021), *WIZ* (cg24203619), and *MGAT3* (cg26348584), among many other loci. Consistent with prior EWAS, we observed strong associations at the *AHRR* locus (cg05575921, cg05934812, cg14817490).

**Figure 1.**
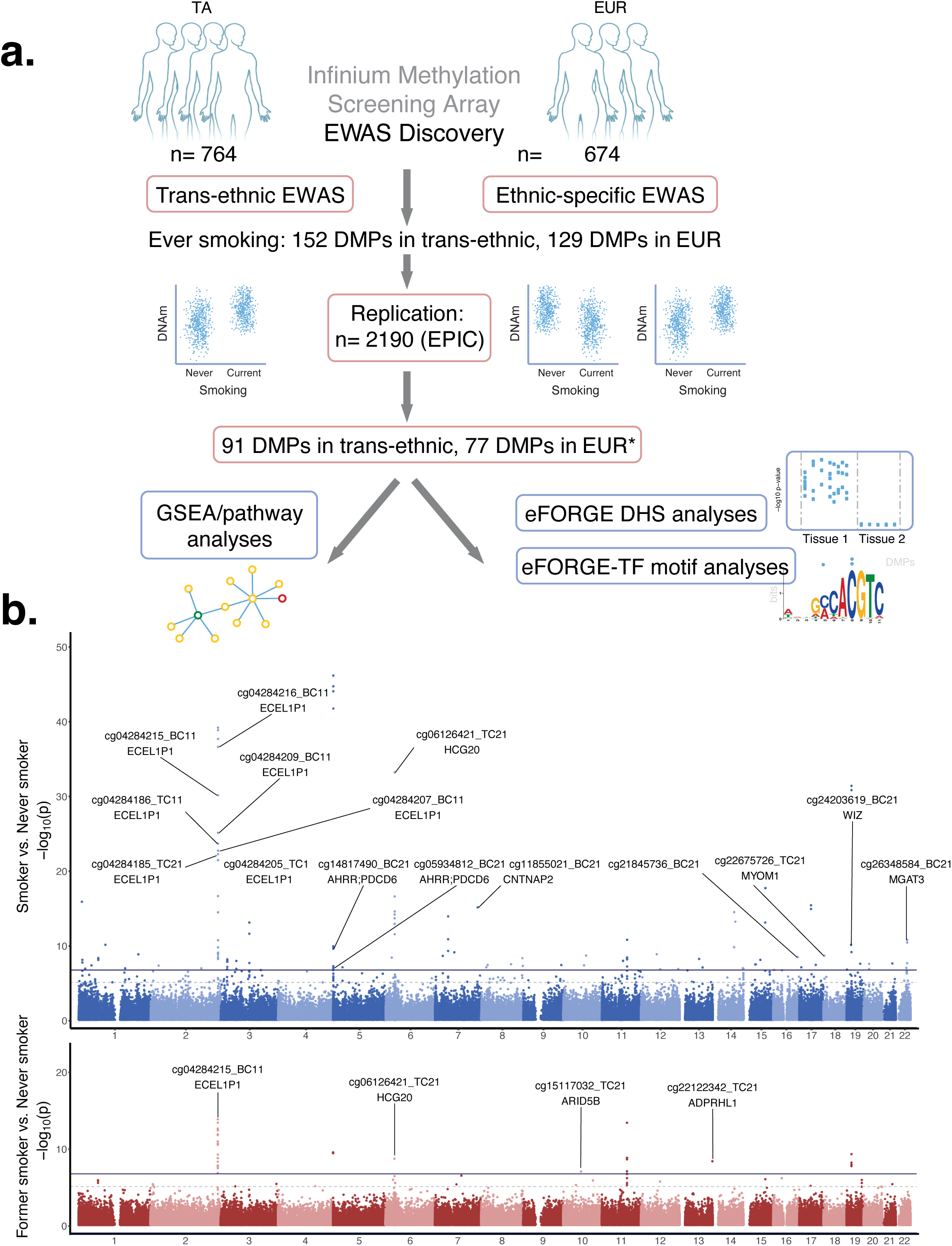
EWAS of smoking using the new MSA platform, which spans 262,470 CpGs: **[a]**Schematic representation of study **[b]** Manhattan plot of novel CpGs and genes in MSA transethnic EWAS (compared to the EPIC array EWAS). Related to **tables S1-S6**. *from Hoang *et al.* 2024, doi: 10.1016/j.ebiom.2023.104956

In comparisons of former versus never smokers, we identified a novel top DMP at the *ECEL1P1* locus (cg04284215), alongside new signals at *ARID5B* (cg15117032) and *ADPRHL1* (cg22122342). In total, we found a range of novel, MSA-specific DMPs, comprising 38 and 44 DMPs in the European (EUR) and trans-ethnic/trans-ancestry (TA) analysis, respectively, with 30 DMPs overlapping between both sets. We also found a range of replicated DMPs for CpGs assayed in the ALHS EPIC array meta-analysis, comprising 83 DMPs in EUR and 97 DMPs in TA. In total, using the MSA we identified 129 DMPs (EUR) and 152 DMPs (TA).

### eFORGE analyses of current vs never smoker DMPs

To further investigate epigenomic signatures of tobacco smoking, we performed integrative analyses across smoking-associated EPIC and MSA DMPs for current vs never smoker using eFORGE, which determines whether the identified DMPs are enriched for active regulatory elements across a wide range of cell types and tissues, thereby providing precise functional context for the observed epigenetic changes^14–16^. Analysis of the top DMPs from the MSA and the ALHS EPIC EWAS meta-analysis revealed significant enrichment of smoking-associated DMPs in blood lineage DNase I hotspots, particularly in HSPCs (q-value < 0.01, top 1000 DMPs ranked by p-value from the MSA EWAS and top 100 DMPs ranked by p-value from the EPIC EWAS, **Figure 2a,b**). These enrichment patterns highlight cell type–specific regulatory roles for identified DMPs in HSPCs.

**Figure 2.**
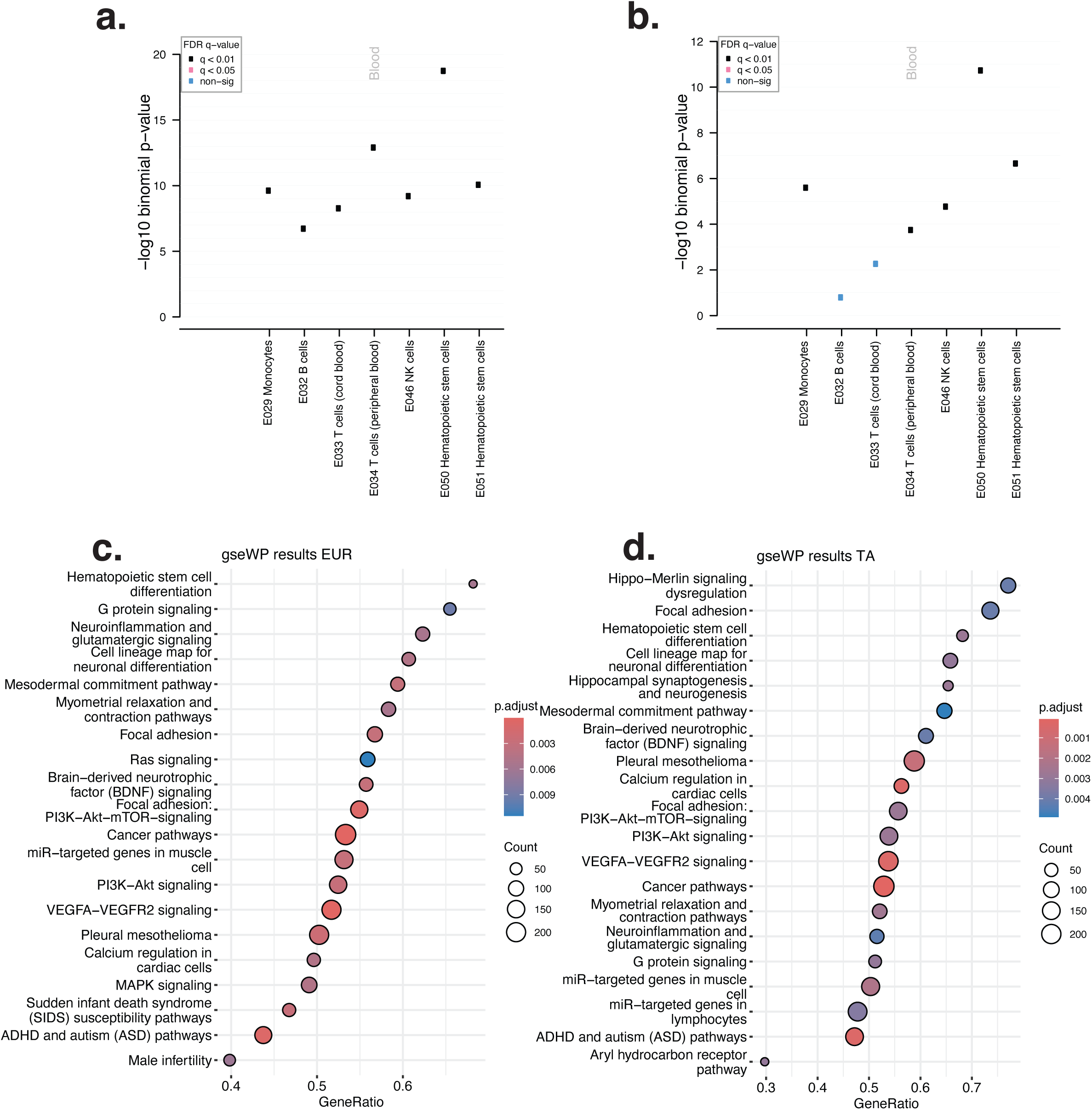
eFORGE and pathway analyses for MSA smoking EWAS: **[a]**eFORGE results for MSA smok-ing EWAS (current vs never smoker), across top 1000 CpGs (tabular results at https://eforge.altiusinsti-tute.org/files/0x6E127A160B7E11F0A42708A13A848967/index.html) **[b]** eFORGE results for EPIC array smoking EWAS (top 100 CpGs, current vs never smoker, tabular results at https://eforge.altiusinsti-tute.org/files/0x951751220B8811F0B39C28AC3A848967/index.html). **[c]** GSEA results for smoking MSA EWAS in Europeans **[d]** GSEA results for transancestry smoking MSA EWAS. Related to online tables https://eforge.altiusinstitute.org/files/0x6E127A160B7E11F0A42708A13A848967/index.html and https://eforge.altiusinstitute.org/files/0x951751220B8811F0B39C28AC3A848967/index.html.

### GSEA analyses of current vs never smoker DMPs

To understand the pathway-level impacts of our epigenomic signatures of tobacco smoking, we performed gene set enrichment analysis (GSEA) on our current vs never smoker EWAS results^17 18^. GSEA of both EUR and TA MSA EWAS results revealed convergent biological themes (80%, 16/20 top-ranked pathways were shared between datasets, **Figure 2c,d**). Prominent among the enriched pathways was “Hematopoietic stem cell differentiation”, which was the top pathway in EUR analysis and the 3^rd^ top pathway in TA analysis. In addition, other pathways were highlighted such as growth signaling pathways (“G protein signaling”, “PI3K Akt mTOR signaling”, “PI3K Akt signaling”), and adhesion pathways (“Focal adhesion”), among others. Taken together with eFORGE results, GSEA findings point to a primary impact of smoking on HSPC regulatory elements and pathways.

### A differentially methylated region at *ECEL1P1* is associated with smoking exposure and maps to a regulatory element

The top DMP for former vs never smoker in our analyses was a novel, MSA-specific DMP at the *ECEL1P1* locus (cg04284215, p<2.17×10^−7^, TA analysis). Further inspection of the *ECEL1P1* locus revealed that this DMP is located within a contiguous differentially methylated region (DMR) spanning 12 adjacent CpGs. Notably, 9 out of 12 DMPs in this DMR are novel to the MSA platform (cg04284215, cg04284209, cg04284171, cg04284205, cg04284186, cg04284185, cg04284170, cg04284216, cg04284207). DNA methylation beta values across these CpGs exhibited a consistent decrease from never to former to current smokers, with cg04284215 showing over 10% change in DNAm (**Figure 3a,b**). Loess curves for beta values at 3 CpGs located at the edge of this DMR showed recovery to average never-smoking baseline levels over time, consistent with prior observations that certain CpG regions—often located at the edges of regulatory elements like enhancers and CpG island shores—exhibit greater DNA methylation variability and dynamic change over time (**Figure 3a**)^19^. When compared with our EPIC EWAS results, novel MSA-specific CpGs at *ECEL1P1* collectively form the most long-lasting and persistent smoking-associated DMR detected so far, with our analyses covering over 60 years since quitting smoking (interquartile range 13.88-34 years)^4,20,21^. In addition, the *ECEL1P1* DMR contains the top DMPs by p-value in our EWAS of former vs. never smoker (**Figure 1b**).

**Figure 3.**
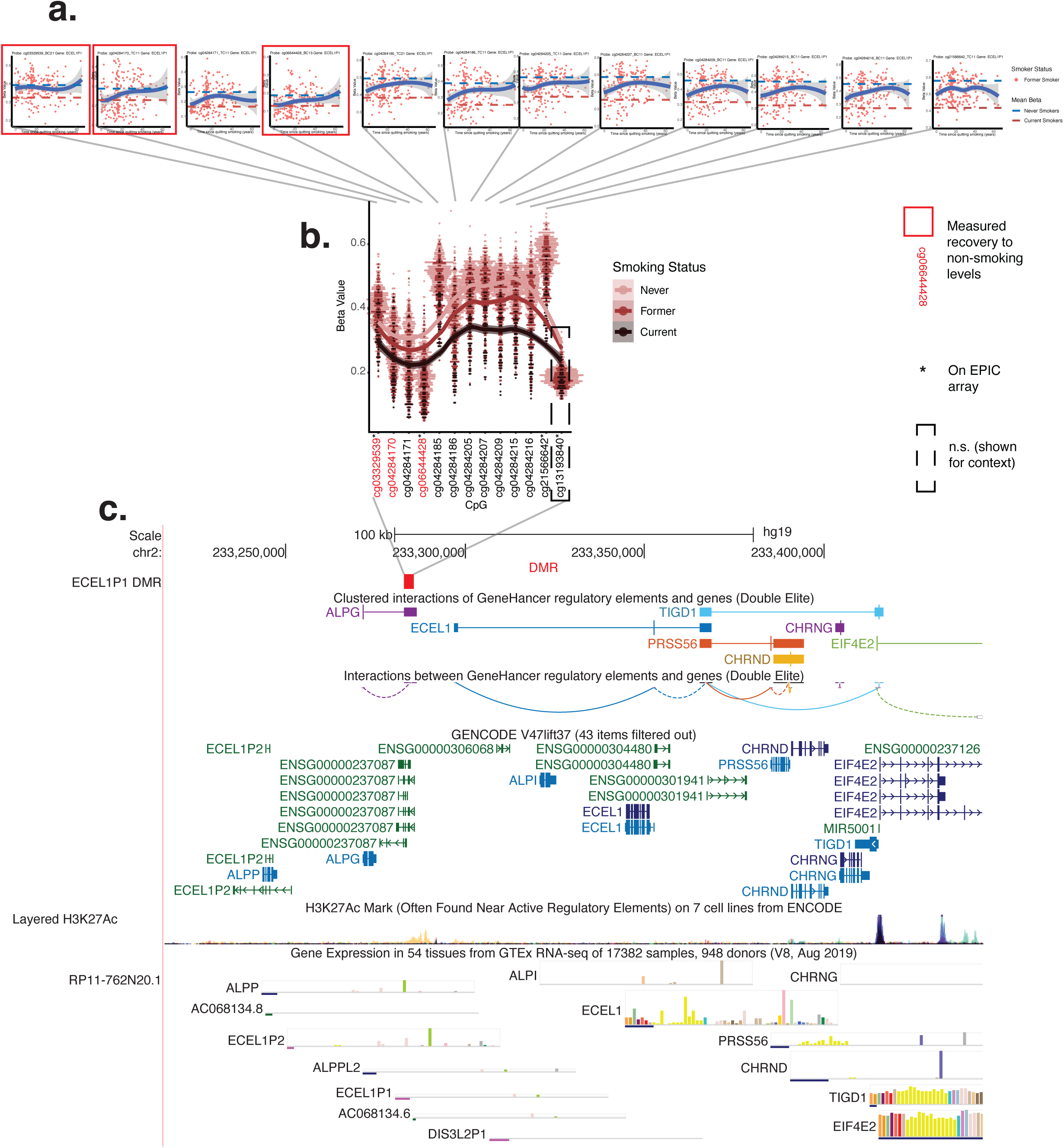
Significant MSA smoking EWAS results at the *ECEL1P1* locus: **[a]**DNA methylation over time at the 12 significant CpGs at *ECEL1P1* (x axis: time since smoking cessation in years, y axis: beta values). Highlighted in red boxes are CpGs with a LOESS curve value that recovers to non-smoking average levels. The three CpGs where recovery to non-smoking average levels is observed are at the periphery of this DMR (CpG IDs highlighted in red in panel b). **[b]** Beta values for never, former and current smokers in CpGs at *ECEL1P1*. These DMPs form a DMR, including novel MSA-specific CpG cg04284215, which is the top DNA methylation signal in former smokers. **[c]** This DMR (shown as a red box) is located in a “Bivalent Enhancer” region in blood that loops to the promoter of *ALPG*. Nearby proximal looping with *CHRND*, which is part of the nicotine receptor, is also shown (GeneHancer track). Related to online table https://eforge.altiusinstitute.org/files/0xE2B6423421-FA11F0A512D79C3A848967/index.html, **figure S1** and **tables S1-S8**.

Further inspection of this locus showed that this DMR overlaps a bivalent enhancer in blood that is proximal to regions with three-dimensional genomic interactions with the promoters of alkaline phosphatase, germ cell (*ALPG*) and cholinergic receptor nicotinic delta subunit (*CHRND*) (GeneHancer interactions, **Figure 3c**)^22,23^. To explore the function of this putative regulatory region, we inspected the role of genetic variation. Through FORGEdb, we identified rs79466634 as a highly prioritized candidate functional variant within this 1117 bp region (FORGEdb score = 10, the maximum score, indicating convergent evidence from eQTLs, ABC interactions, TF motifs, DNase I hotspots and histone marks)^24,25^. AlphaGenome analysis of rs79466634 showed that alleles of this variant had predicted effects on transcription of lncRNA ENSG00000237087 in *cis* across HSPCs and T cells^26^ (**Figure S1**). Further, rs79466634 is an eQTL for *EIF4E2*, is associated with *CHRNG* through ABC^27^, is located in an ENCODE4 regulatory element, and is predicted to impact multiple TF motifs (https://forgedb.cancer.gov/explore?rsid=rs79466634). Taken together, data suggest that this is a regulatory element with predicted effects on transcription in *cis* and predicted associations with *EIF4E2, CHRNG* and lncRNA ENSG00000237087, among other genes —such as previous reports on *ALPP*^28^— warranting further investigation. These results additionally suggest a potential relationship between smoking-associated DNAm changes and the regulation of pluripotency and nicotinic acetylcholine receptors.

### Cell type–specific DNAm changes highlight neutrophil-specific *AHRR* hypomethylation

To analyze smoking-associated DNAm shifts at the level of individual blood cell types, we analyzed our EWAS with CellDMC integrated with EpiDISH cell type proportion estimates^6,29,30^. First, we evaluated associations between blood cell type proportions and smoking (adjusting for the same factors as in EWAS, **Methods**), identifying a significant association between monocyte proportions and smoking status (p<0.05). We then proceeded to evaluate cell type-specific differentially methylated CpGs (differentially methylated cytosines in individual cell types or DMCTs). DMCTs for current vs. never smoker were almost exclusively observed in myeloid cells, specifically neutrophils. Nearly all of the coefficients were negative, indicating reduced methylation in current smokers. Far fewer results were observed for former vs. never smoker, including one DMCT observed in CD4 T cells (**Figure 4a, Tables S9-S15**). The canonical smoking marker CpG cg05575921 displayed pronounced neutrophil-specific DNA hypomethylation in current smokers (but not former smokers, **Figure 4b**). This DMP maps to an H3K27Ac-marked enhancer at the *AHRR* locus (**Figure 4c**), suggesting a mechanistic link between tobacco smoke exposure and myeloid-specific epigenetic regulation at *AHRR*, which is short-lived and reversible within 1 year of smoking cessation, as shown by the lack of association in former smokers with 1 year or more since quitting smoking. These results show that a subset of “current vs never smoker” and “former vs never smoker”-associated DMPs are cell type-specific, mostly occurring in neutrophils and typically representing more short-lived and reversible DNAm changes.

**Figure 4.**
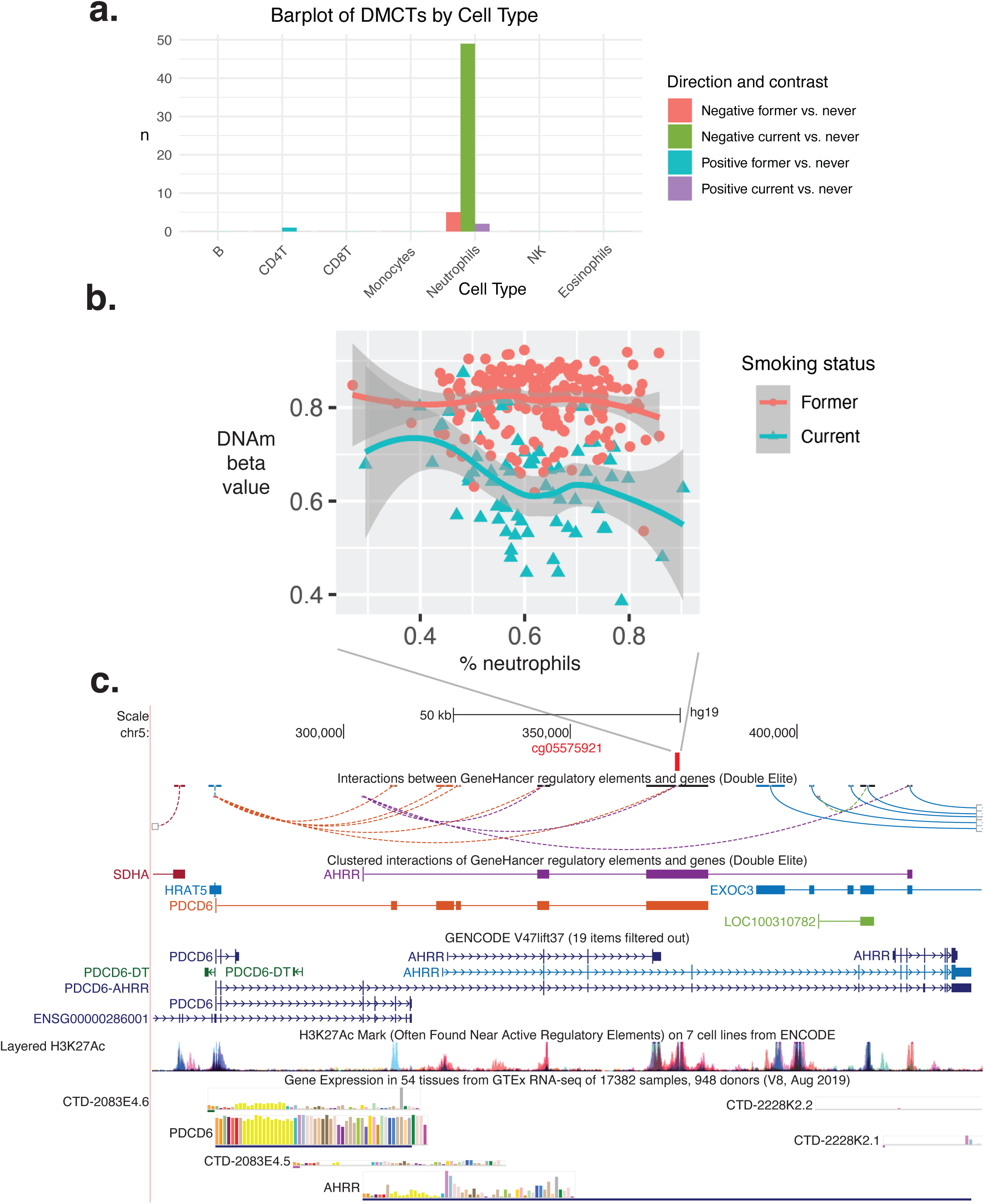
Cell type-specific DNA methylation in MSA smoking EWAS: **[a]**CellDMC results for MSA smoking EWAS, across different EpiDISH blood cell types. In agreement with previous analyses (You *et al.*, 2020, Nat Comms), we saw the largest number of DMCTs in myeloid cells, specifically neutrophils. **[b]** A top DMCT for smoking is at cg05575921, which presents lower DNA methylation in current smokers within neutrophils. **[c]** cg05575921 is located at the *AHRR* locus. Specifically cg05575921 is in an eFORGE enhancer region in blood that loops to the promoter of *PDCD6* and to *AHRR* (CpG shown in red, see https://eforge.altiusinstitute.org/-files/0xAA37E37620B611F0A7472C0E3B848967/index.html for enhancer data). Related to tables **S9-S15**.

### An MSA-specific differentially methylated position at *RUNX1* links hematopoiesis to smoking exposure and maps to a regulatory hub

We also sought to characterize smoking-associated DMPs directly impacting HSPC pathways, as suggested by our GSEA analysis. Among our top novel MSA-specific CpGs is a prominent smoking-associated DMP, cg25815220, within the *RUNX1* locus—an essential regulator of HSPC generation and differentiation. DNAm at cg25815220 showed a graded response to smoking status, with the lowest levels in current smokers, intermediate levels in former smokers, and highest levels in never smokers (**Figure 5a**). This CpG lies within an intronic regulatory region of *RUNX1* marked by a genic enhancer chromatin state in HSPCs, hinting towards potential functional impact of this region as a genic enhancer for *RUNX1* (**Figure 5b, Files S1-S2**). Persistent low DNAm levels in former smokers over 1 year after cessation of smoking suggest that epigenetic memory at this enhancer is retained over 1 year. HSPCs, unlike most blood cell types, are long-lasting, with individual HSPC clones living up to almost 60 months^31^. HSPCs are the only plausible cell population capable of registering this signature over this timeframe and propagating this signature across different blood cell populations^32^. When exploring chromatin looping data, we noted an absence of *RUNX1*-linked chromatin conformation data from primary HSPCs in GeneHancer data. This likely reflects the scarcity of this primary cell type, which can be harder to obtain than peripheral blood lymphocytes and is often underrepresented in global genomics consortia^33^. Altered DNAm at the *RUNX1* locus, taken together with results for GSEA and eFORGE, as well as other EWAS DMPs, highlight HSPC CpGs and HSPC-regulating pathways as central to the impact and persistence of smoking-associated differential DNA methylation.

**Figure 5.**
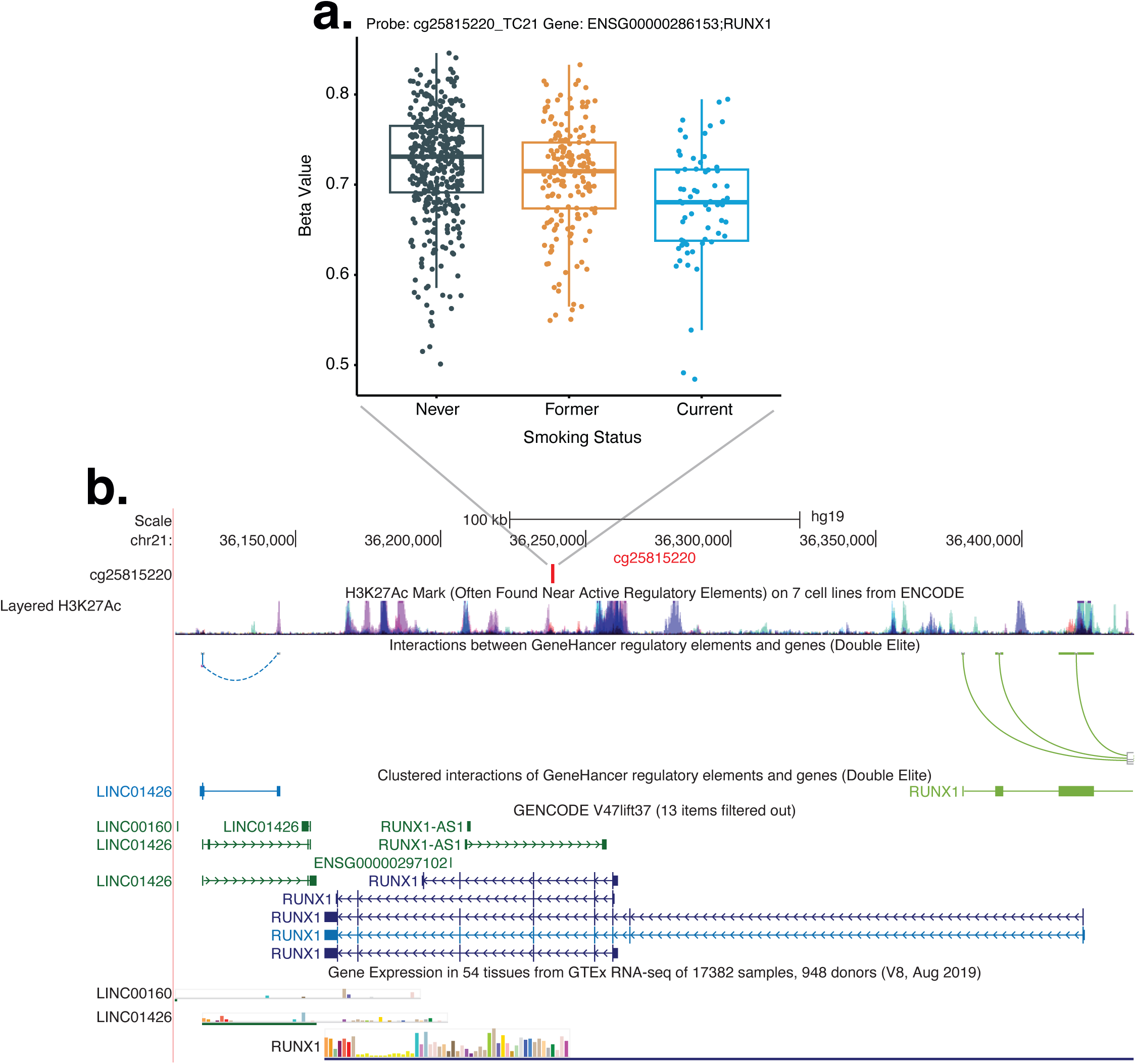
Significant MSA smoking EWAS results at *RUNX1*, a major HSPC regulator: **[a]**beta values for never, former and current smokers in novel MSA CpG cg25815220 at *RUNX1*. **[b]** This DMP (shown in red) is located in an intronic regulatory region within *RUNX1*, marked by DNase I hotspots across several Roadmap Epigenomics tissues (**file S2**) and “genic enhancer” chromatin state in several Roadmap Epigenomics tissues (**file S3**). Note that, even in former smokers (ceased >1 year before sample obtention) have lower DNAm at this *RUNX1* enhancer. The only cell type with a lifespan >1 year that could register this long-term DNAm signature and then perpetuate it across descen-dant blood cell types is that of hematopoietic stem and progenitor cells (HSPC), which might also explain the lack of 3D genomics data, as HSPCs are hard to obtain in large numbers as primary cells, and are often lacking from genom-ic assays that require high amounts of DNA. Related to **tables S1-S6** and **files S1-S2**.

### Cell type-specific and shared hematopoietic DNAm changes in smokers

Overall, a clearer picture of smoking-associated DNA methylation emerges. Analyses of these data considering time windows involved (e.g. former smoker over 1 year after quitting smoking), cell type-specificity, pathway information and functional annotations revealed two broad classes of DNA methylation changes in smoking (**Figure 6**). First, transient, cell type–specific changes were observed primarily in neutrophils/myeloid cells (**Figure 6a**), representing mostly temporary, reversible responses to current smoking exposure. In addition, we observed a second class of alterations spanning multiple hematopoietic lineages and likely originating in HSPCs, manifesting as persistent, “memory”-like long-lasting DNAm patterns in former smokers (**Figure 6b**). These changes were associated with dysregulated hematopoiesis and implicated in processes such as growth pathways, with specific genes involved in erythropoiesis, transposon repression, and genomic stability, for example *RUNX1, DMTN, HNRNPA1,* and *XRCC3* (**Figure 2c,d**, **Tables S1-S8**). These enduring, persistent DNAm changes may act as molecular intermediates linking smoking to increased risk for hematologic and systemic disease. Taken together, our combined results revealed a dual pattern of smoking-induced epigenetic remodeling—comprising both transient, blood cell-type-specific signatures and long-lasting, HSPC-derived changes—offering a molecular framework potentially underlying long-term immune-associated effects^34^ and providing mechanistic insight into smoking-associated disease susceptibility. Collectively, these results support a model where tobacco smoking drives both short-term and persistent epigenetic changes in blood, with long-term mechanistic links to hematopoietic dysregulation. Persistent DNA methylation changes at HSPC regulatory regions in key loci, such as those at the *ECEL1P1* locus, provide a plausible mechanistic substrate for long-term, residual disease risk among former smokers with many years of abstinence.

**Figure 6.**
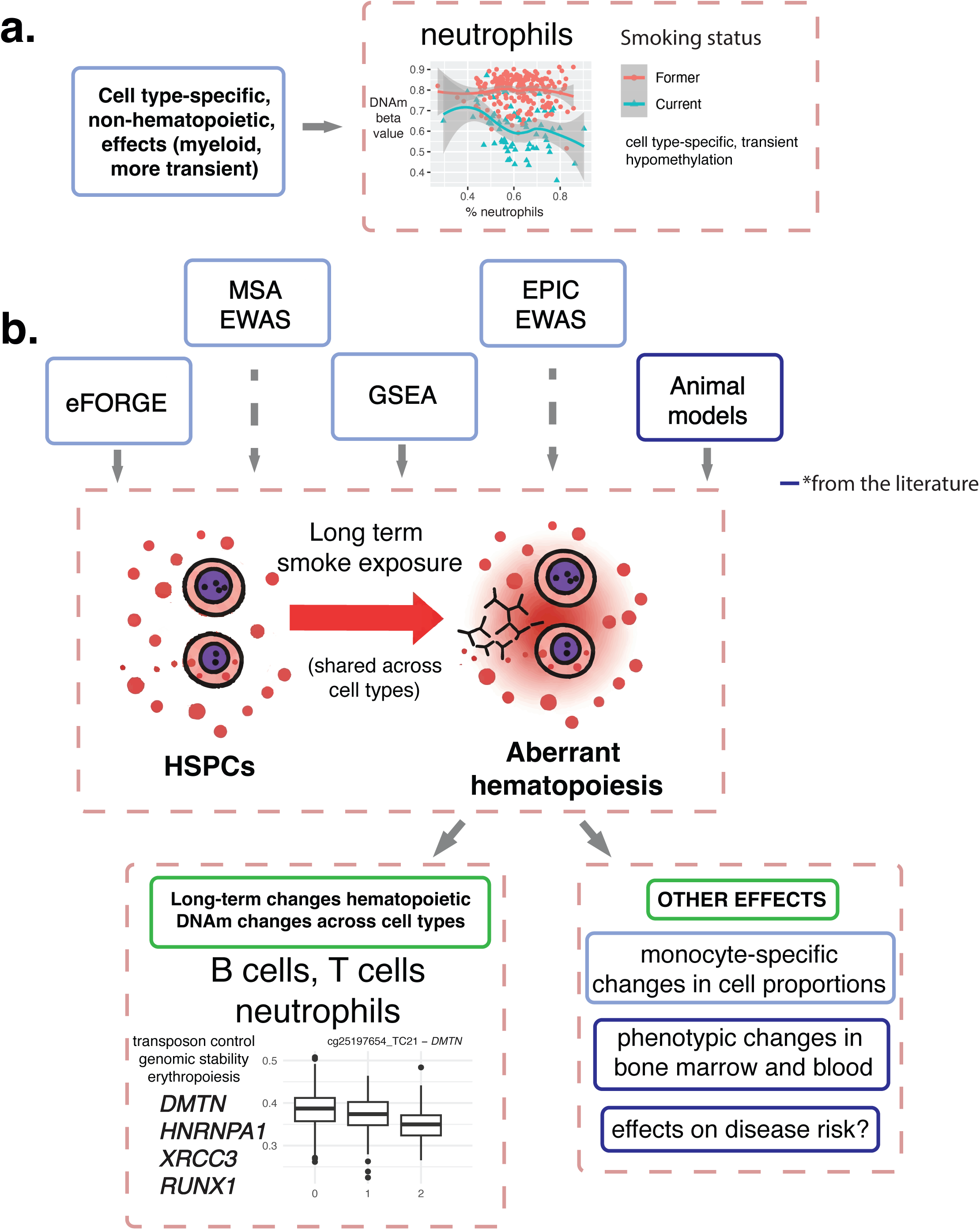
Cell type-specific and shared hematopoietic DNAm changes in smoking: **[a]**Schematic representation of cell type specific DNAm changes in smoking, which are more transient and restricted to myeloid cells (neutrophils) **[b]** Schematic representation of DNAm changes shared across different blood cell types in smoking, which originate in hematopoietic stem cells and broadly outlast the lifespan of most/all peripheral blood cells, constituting a retained memory of smoking-related aberrant hematopoiesis.

## Discussion

Among all exposures investigated via EWAS, tobacco smoking consistently emerges as one of the strongest factors associated with DNAm changes. Previous studies have reported a range of DNAm alterations in smoking, with top DMPs at *AHRR* and *F2RL3,* among many others. Some of the observed DMPs persist for several years after smoking cessation^3^. DNAm changes affect numerous genes across multiple tissues, with considerable overlap in both the direction and genomic location of the DNAm changes across tissues^1^. Smoking effects on DNAm can be detected at the cell type-specific level via cellDMC, with published reports highlighting robust and reproducible associations between smoking and cell-specific methylation changes^5,7^.

Despite these advances, improved understanding of the underlying biology of smoking-associated DNAm changes is needed to inform future risk stratification, individualized smoking cessation programs, as well as prevention and treatment strategies for smoking-driven chronic or noncommunicable diseases^35–38^. EWAS reveal a predominantly myeloid bias in the cell-specific effects of smoking^7^. In addition, several studies report an increase in monocyte proportions in the peripheral blood of smokers^8,9^. To better understand the underlying biology of smoking-associated DNAm changes, additional knowledge of the cell type specificity and cellular lineage of smoking-associated DNAm is required.

In this study, we identified a broad range of novel cell type-shared and cell type-specific DMPs and genes associated with smoking, as well as confirming previous observations, such as a reported increase in monocyte cell proportions. Crucially, we systematically integrate these results with their broader regulatory context. Our analyses show that the long-term DNAm signature induced by smoking is at least in part anchored in altered regulatory elements within HSPCs, providing a biological substrate for the propagation of an epigenetic memory that endures for years. This aberrant hematopoietic regulation in smokers appears to comprise a range of different processes, including anomalous DNAm patterns at major HSPC regulators such as *RUNX1*, alterations in blood cell proportions—consistent with previously reported increases in monocytes, which we also find—and altered DNAm patterns at a range of additional genes involved in erythropoiesis (e.g. *DMTN*), transposon control (*HNRNPA1*)^39^ and genomic stability and repair (*XRCC3*)^40^ (**Tables S1-S8**).

A key finding of our study is that the most enriched cell type in our eFORGE analysis are CD34 positive HSPCs, indicating that among the regulatory regions assessed by chromatin accessibility assays, HSPCs provide the strongest signal for the smoking-associated DMPs. This observation highlights a potential role for HSPCs as candidates for propagating the epigenetic effects of smoking. Furthermore, GSEA revealed hematopoiesis as one of the top 3 categories in both our EUR and TA datasets, with critical hematopoietic regulators present such as *CD34, RUNX1*, and *GATA2*. Interestingly, canonical epigenetic editors with roles in HSPCs such as *DNMT3A* did not emerge as significant, which may be attributable to our younger study population, a group less likely to exhibit clonal hematopoiesis.

The aforementioned DMPs at an enhancer within *RUNX1—*which orchestrates lineage commitment between myeloid and lymphoid compartments—are especially interesting given the observation of a shift in myeloid/lymphoid cell proportions in smokers, which we and others have observed^7–9^. Whether the myeloid/lymphoid cell proportion shift is orchestrated by *RUNX1* itself or by other regulators within a smoking-associated aberrant hematopoietic compartment^41^ warrants further investigation.

One of the limitations of this study is the use of DNA methylation derived from peripheral blood, which is common in EWAS and large-scale molecular epidemiology analyses. Future studies may seek to expand our findings focusing on the bone marrow niche where hematopoietic stem and progenitor cells reside. In addition, while we employed rigorous cell-type proportion adjustments across B cells, NK cells, CD4 and CD8 T cells, monocytes, neutrophils and eosinophils to account for blood composition heterogeneity, current bulk tissue EWAS may lack the sensitivity to detect epigenetic shifts occurring within specific, rare leukocyte subpopulations.

Building upon a comprehensive comparison with prior EPIC array EWAS data, our study demonstrates that while several of the identified genes have prior associations with tobacco exposure, the expanded genomic coverage of the MSA was influential in refining our understanding of the underlying biology. By assaying over 123,000 CpGs not previously studied, we identified 61 novel smoking-associated DMPs, including entirely new loci at *CNTNAP2*, *WIZ*, and *MGAT3*, and also at known genes like *AHRR*. Most notably, this enhanced resolution allowed us to characterize DNAm changes spanning 12 CpGs in a 1117 bp region at *ECEL1P1* as the most long-lasting, persistent smoking-associated DMR detected to date, a discovery that underscores the MSA’s unique capacity to map stable, HSPC-derived DNA methylation changes that were largely invisible to previous arrays.

We also observed cell type-specific differentially methylated CpGs (DMCTs) in neutrophils, confirming previous reports^7^. These DMCTs occur predominantly at myeloid/granulocyte regulatory elements. Globally, these changes seem to be transient and are mainly detected in current vs never smokers. This is consistent with the lifespan of neutrophils, which are a short-lived cell type^42^. We also find many DNAm changes shared across blood cell types, including a DMR at *ECEL1P1*, the most long-lasting, persistent smoking-associated DMR detected so far. This *ECEL1P1* DMR may prove useful for the detection of former smokers and the modelling of smoking-associated disease risk. Taken together, these data suggest that the DNAm effects of smoking can be broadly categorized into two domains. The first involves more transient, cell type–specific changes, predominantly affecting myeloid cells which are likely responding to smoking exposure in peripheral blood. The second domain comprises long-term changes that are shared across cell types, likely resulting from an aberrant hematopoietic compartment and the transmission of an epigenetic memory of smoking within HSPCs.

## Conclusion

In conclusion, our study expands the mapping and biological understanding of the DNAm effects of smoking and contextualizes them within a dual framework of short-term, cell type–specific responses and long-term hematopoietic regulation. This integrated view not only corroborates previous literature but also provides new insights into the mechanistic underpinnings of smoking-induced epigenetic alterations.

## Acknowledgements

Illumina provided the MSA laboratory analyses at no cost as part of the MSA early access program. The current work was supported by intramural funding from the National Cancer Institute (Z01-CP010119), National Institute of Environmental Health Sciences (Z01-ES102385, Z01-ES049030, Z01-ES043012, and contract number HHSN273201600003I), and National Heart Lung and Blood Institute (1ZIAHL006285-02). The ALHS was also supported in part by American Recovery and Reinvestment Act (ARRA) funds through NIEHS contract number N01-ES-55546. This work utilized the computational resources of the NIH HPC Biowulf cluster (https://hpc.nih.gov). We thank the NIEHS Molecular Genomics Core Laboratory (Kevin Gerrish) for technical support. We thank Michael Shtutman for his insightful comments regarding AlphaGenome analysis. This research was supported by the Intramural Research Program of the National Institutes of Health (NIH). The contributions of the NIH authors are considered Works of the United States Government. The findings and conclusions presented in this paper are those of the authors and do not necessarily reflect the views of the NIH or the U.S. Department of Health and Human Services.

## Data availability

The data underlying this investigation will be provided upon request as described on the AHS website: https://aghealth.nih.gov/collaboration/studies.html.

## Supplementary information

Supplementary tables are available from https://drive.google.com/drive/folders/13a-3-rROYnPmFeXfRGlIMezGuEwiJPNQ?usp=sharing

## Materials and Methods

### 1. Participants

The Agricultural Lung Health Study (ALHS) has been described in detail^43,44^. Briefly, ALHS is a nested case-control study of current asthma within the Agricultural Health Study (AHS)^45^, a prospective cohort study of 89,654 farmers and their spouses from Iowa and North Carolina, USA. The ALHS sub study included over 3301 participants (1223 current asthma cases; 2078 noncases) from the AHS enrolled between 2009 and 2013 based on responses to an AHS questionnaire administered in the period 2005–2010. Participants self-reported their smoking history via questionnaires administered at the various AHS survey waves and at enrollment in ALHS and were categorized as never smokers (person who has smoked <100 cigarettes in his/her lifetime), former smokers, current smokers at the time of ALHS enrollment, or missing assessment. Information on exposure to secondhand smoke was also recorded and defined as passive smoke exposure at home or smoke exposure for at least one hour per day on average.

The ALHS enrolled asthma cases using three criteria: a) having self-reported current diagnosed asthma without any self-reported diagnosis of chronic obstructive pulmonary disease (COPD) or emphysema (n=876); b) having current self-reported diagnosed asthma with COPD or emphysema diagnosis among never smokers or light, former smokers (≤10 pack-years) (n=38); or c) having potential undiagnosed asthma identified by self-report of current asthma symptoms or use of asthma medication without COPD or emphysema diagnosis among never smokers or light, former smokers (≤10 pack-years) (n=309). Participants free of asthma based on the above criteria (n=2078) were randomly selected from the AHS participants without the above criteria.

DNA was extracted from peripheral blood samples collected from the participants at ALHS enrollment^46^. In a subset of 887 participants from ALHS (239 current asthma cases; 648 noncases), the DNA samples were bisulfite treated using EZ-96 DNA Methylation kits (Zymo Research Corp.) and scanned using the Infinium Methylation Screening Array (MSA) as per manufacturer’s protocols^13^. Selection of the 887 participants for the MSA scan was performed using the following criteria: (a) availability of extracted DNA without a “No Future Use” restriction flag, and (b) absence of a previously failed exome chip assay indicating poor biospecimen quality, yield, or contamination. Among eligible participants, all 787 individuals without prior DNAm data on the Illumina EPIC or 450K platforms were included. An additional 100 individuals (50 current smokers and 50 non-smokers, randomly selected) with prior EPIC methylation data were also included to enable direct cross-platform comparison. All participants provided informed consent. The Institutional Review Board at the National Institutes of Health approved this study. For our analyses we used AHS data version P3REL201209.00.

### 2. DNA methylation pre-processing, quality control, and cell-type proportion estimation

Of the 887 participants who were scanned with the MSA, we conducted analyses on 764 participants (186 current asthma cases; 578 controls) after quality control, removal of outlier samples, and retaining samples that also had measured genotype data. Sensible Step-wise Analysis of DNA Methylation BeadChips (SeSAMe) was used for quality control and normalization^47^. Briefly, the infer channel algorithm was applied to Infinium I probes to determine the appropriate color channel. Non-linear dye-bias correction was performed to adjust for intensity differences between dye channels. Detection p-values were calculated using <u>p</u>-value with OOB probes for <u>A</u>rray <u>H</u>ybridization (pOOBAH), and background subtraction was implemented using noob, normal-exponential deconvolution using out-of-band probes. Beta distributions of Infinium I and Infinium II probes were matched to correct for probe-type–specific biases. Probes overlapping SNPs, cross hybridizing probes, probes on sex chromosomes, CH probes, and probes having a detection p-value >0.05 in >50% of samples were excluded. DNAm was measured using noob-background corrected^47,48^, normalized beta values ranging from 0 to 1, representing 0-100% methylation. Samples with a sex-mismatch (n=40), missing genotype data (n=5), missing >10% probes (n=78), or identified as normalized-beta outliers were excluded from analysis. A total of 230,325 probes in 764 samples were used to conduct analysis following quality control measures. Missing DNAm values were imputed using K-Nearest Neighbors with a k=10. Singular value decomposition (SVD) plots were used to view sources of technical variation. ComBat, implemented in ChAMP, was then used to correct for batch and technical effects: sentrix ID, sentrix position, and sample box^49^.

#### Estimated Immune Cell-Type Proportions

Seven immune cell-type proportions (B cells, NK cells, CD4 and CD8 T cells, monocytes, neutrophils and eosinophils) were estimated using EpiDISH on ComBat adjusted beta values, a framework for reference-based inference of cell-types using DNase I Hypersensitive Site specific information for deconvolution of intra-sample heterogeneity as previously described^29,30^. Using the cell fractions produced by EpiDISH, cell-type specific changes in methylation between never and current smokers, never and former smokers, and former and current smokers were identified using CellDMC^6^.

### 3. Genotype processing

Details of the ALHS’s genetic data and genotyping procedures have been published elsewhere ^50,51^. Participants were genotyped using the UK Biobank Axiom Array (Axiom_UKB_WCSG) by Affymetrix Axiom Genotyping Services (Affymetrix, Inc., Santa Clara, CA). Calls were extracted using Affymetrix Power Tools. Following extraction, variants with a missing rate > 5% or a Hardy-Weinberg p-value < 1×10^−6^, or MAF < 0.05 were excluded. After quality control, genetic principal components were generated using PLINK v1.9.^52^

### 4. Statistical analysis

Epigenome-wide analysis of DNAm was contrasted for current versus never smokers, as well as former versus never smokers, using a survey-weighted general linear regression to account for asthma status in the nested case-control study. Models were adjusted for asthma status, sex (male and female), age at home visit (years), body mass index (BMI; kg/m^2^), secondhand smoke exposure (yes and no), residing state (Iowa or North Carolina), race and ethnicity (to account for shared environment, dietary, and social determinants of health), the first 10 genetic principal components generated from genotype data (to account for genetic ancestry), and 7 immune cell subtype estimations from EpiDISH (B cells, NK cells, CD4 and CD8 T cells, monocytes, neutrophils and eosinophils). Analyses were also stratified by EUR and TA populations. Details of additional stratified and unweighted sensitivity analyses can be found in the *Supplemental Materials* (**Tables S1-S8**). For example, we reran the EWAS excluding 2 former smokers who quit smoking within one year of blood draw but this did not affect findings (**Tables S7-S8**). Stratified and unweighted analyses can be found in the Supplemental Materials. Linear models employed to explore the association between cell proportions and smoking status performed adjustment for the same variables as our EWAS model. Statistical analyses and plots were generated with R statistical software (version 4.3.1)^53^ using ggplot2^54^, cowplot^55^, and a modified version of miamiplot^56^.

### 5. CpG probe annotation and filtering

CpG probes were annotated using the Illumina MSA manifest^13,57^ and Zhou et al. GENCODEv41 gene annotation file^57^. Probes were filtered for epigenome-wide significance at a Bonferroni adjusted p-value ≤ 2.17 × 10^−7^ (0.05/230,325 probes). Probes meeting significance were included in downstream analyses.

### 6. Enrichment of genomic features

We performed an eFORGE v2^14–16^ analysis on top 1000 most significant CpGs from the MSA EWAS using standard settings to identify tissue or cell-type specific signal. For further functional enrichment and biological insight, we performed GSEA on the full distribution of CpG probes using clusterProfiler^58,59^ and wikiPathways^60^. We included pathways with a nominal p-value < 0.05.

---

**Figure S1:**
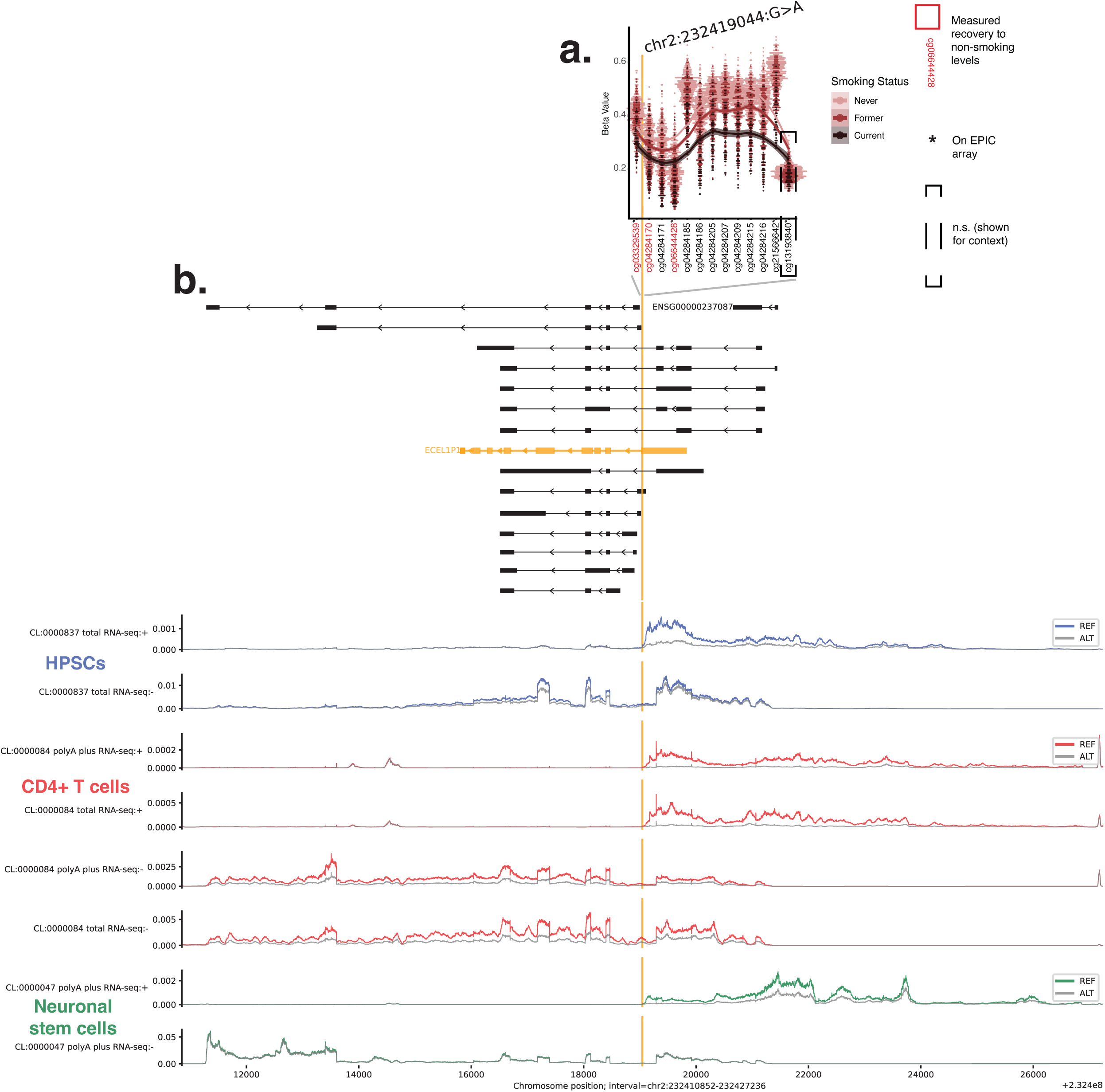
Cell-type-specific effects of DMR-overlapping variant rs79466634 on lncRNA ENSG00000237087 across cell type lineages predicted by AlphaGenome: **[a]**Beta values for never, former and current smokers in CpGs at *ECEL1P1*. These DMPs form a DMR, including novel MSA-specific CpG cg04284215, which is the top DNA methylation signal in former smokers. Variant rs79466634 is located between DMR CpGs cg03329539 and cg04284170. n.s. (not significant). **[b]** Gene tracks and comparative RNA-seq tracks for the rs79466634 variant (chr2:232,419,044:G>A) across three human cell lineages: hematopoietic stem cells (HPSCs, CL:0000837; top), CD4+ T cells (CL:0000084; middle), and neuronal stem cells (NSCs, CL:0000047; bottom). RNA-seq read density (normalized counts) is shown for the reference (REF, G allele; grey) and alternate (ALT, A allele; coloured) genotypes. Strand-specific expression profiles reveal a stronger transcriptional effect on the positive (+) strand in HPSCs, where the REF (G) allele is associated with an increase in transcript abundance for the long non-coding RNA ENSG00000237087. Notably, similar regulatory effects are observed in both the HPSC and T-cell lineages, suggesting a potential shared regulatory mechanism. In contrast, NSCs show different profiles, highlighting potential lineage-restricted activity of the underlying regulatory element. Genomic context tracks (top) indicate ENSG00000237087 transcript isoforms (in black) and the neighboring *ECEL1P1* pseudogene (in orange). The vertical orange line indicates the position of the candidate causal variant, rs79466634, localized within the bivalent enhancer/TSS flanking region of the affected lncRNA. Interval shown: chr2:232,410,852– 232,427,236 (hg38).

